# Large language models for cancer registry abstraction: a real-world evaluation across models, variables, and cancer types

**DOI:** 10.64898/2026.06.25.26356626

**Authors:** Joshua T Fuchs, Matthew J Satusky, Peter J Leese, Subhadeep Nag, Isaiah W Zipple, Chris D Baggett, Sydney Lash, Katherine Reeder-Hayes, William A Wood, Cara T Johnson, Claire Critchley, Ashok K Krishnamurthy, Jennifer Elston Lafata, Caroline A Thompson, Melissa A Troester, Emily R Pfaff

## Abstract

Cancer registries enable cancer surveillance at the population level. These registries require significant human-time to read through many different parts of the electronic health record, including structured data and lengthy, free-text clinical reports, to abstract values for hundreds of required variables. Large language models (LLMs) offer the possibility to significantly improve this process by supporting and speeding up cancer registry data abstraction. However, it is unclear how well these models perform at real-world cancer registry abstraction involving multiple cancer types and large patient volumes. Here, we evaluate five foundational LLMs for their ability to reliably abstract cancer registry variables. We leverage hospital cancer registry data from a large regional health system as the ground truth and use LLMs to abstract from clinical reports eight registry variables for 5,939 patients with seven different cancer types. We use a zero-shot prompting strategy to compare LLM ability on commonly abstracted cancer variables with different data types. The results show that larger and more advanced models (Claude Sonnet 4.5, GPT-OSS-120b, GPT-OSS-20b) generally outperform smaller models (Gemma 12b, LLaMA 3.1 8b). The best performing models show F1 scores around 0.8 for cancer registry variables with low cardinality (grade, summary stage, laterality), with only slightly lower F1 scores for variables with high cardinality (primary site, regional nodes examined, regional nodes positive). On the more complex task of precise date extraction, all models showed decreased performance on both diagnosis and treatment dates (exact accuracy ∼0.55 for the best performing models), which increased to ∼0.85 for a tolerance within ±30 days. These results quantify the performance of various models as well as the potential and limitations of LLMs in cancer registry abstraction tasks.

**Author Summary:** Cancer registries help us understand who gets cancer, how it is treated, and how outcomes change over time. Today, trained registrars read many parts of the medical record and manually enter hundreds of details, leading to significant delays before cancer researchers can use this data. We tested whether Large Language Models could help with this work. Using clinical reports from 5,939 patients across seven cancer types at a large regional comprehensive cancer center, we asked five different models to capture key cancer registry fields such as where the cancer started, how advanced it is, the tumor’s grade, and the date of diagnosis. In our study, larger models were generally more accurate than smaller ones, and some open-source models performed comparably to proprietary options. Dates were the hardest information to capture precisely. We provide a realistic benchmark for speeding registry workflows in consultation with cancer registrars.

## Introduction

Cancer registration involves state and federal mandated processes for the identification, data collation, and reporting of cancer cases through a network of hospital cancer registrars. These registries form the foundation of population-level cancer surveillance in the United States [1] and enable health systems, researchers, and policymakers to understand cancer incidence, treatment patterns, and long-term outcomes across diverse patient populations. Typically, cancer registrars manually abstract detailed clinical information from the electronic health record (EHR) and enter structured values (e.g., cancer type, diagnosis date, histology, treatment information) into standards such as the North American Association of Central Cancer Registries (NAACCR) data model. While this data collection is valuable and necessary for high-quality cancer research, manual population of these registries is meticulous, expertise-dependent, and time-consuming, often requiring careful human review of pathology reports, operative notes, radiology interpretations, and other clinical documentation.

Despite its essential role in cancer control, manual abstraction remains a significant obstacle, where local registrars are processing hundreds of cases a year, each taking multiple hours[2], resulting in delays for use of these data for trial recruitment and care optimization. Further, it can take up to three years for the aggregated, research-ready data to become available from the central state registry [3]. This delay makes time-sensitive applications such as clinical trial identification, cohort discovery, and policy development and analysis substantially more challenging or impossible.

Large Language Models (LLMs) offer a compelling opportunity to transform the cancer registry workflow. These models have demonstrated an ability to interpret unstructured clinical text and produce structured outputs, even with minimal task-specific training [4,5]. Previous work on transformer architectures and efficient attention mechanisms [6,7] showed that language models can generalize new tasks through few-shot or zero-shot prompting. More recently, the release of frontier-scale models such as GPT-OSS-120B and GPT-OSS-20B [8], Llama-3.1[9], and proprietary models such as Claude Sonnet 4.5 have demonstrated high levels of accuracy in reasoning, summarization, and domain-specific extraction. Several recent reviews and benchmark studies have highlighted the growing applicability of LLMs in clinical information extraction, clinical decision support, and biomedical question answering [10,11]. Other investigations have focused on assessing LLMs’ confidence calibration, error profiles, and sensitivity to complex clinical nuances [12]. This indicates that LLMs are starting to play a meaningful role in biomedical information extraction.

Within oncology, traditional natural language processing systems have historically struggled with essential registry fields due to the highly variable ways clinicians document cancer information [13–15]. Several studies have examined LLMs for tasks such as date extraction or patient-facing summarizations [16,17]. Others have demonstrated success using LLMs for classifying cancer cases or extracting cancer registry information [18–22], but have been restricted to narrow use cases such as single cancer types, limited report types, a limited subset of registry variables, and/or small datasets. These findings suggest that LLMs can reduce the burden of manual cancer data abstraction and enable scalable extraction pipelines. However, the field still lacks a systematic evaluation of how modern LLMs perform at a representative, real-world cancer registry abstraction task involving multiple cancer types, large patient volumes, and a standardized set of multiple cancer registry variables.

In this study, we compare the cancer registry extraction performance of five state-of-the-art (as of 2025) LLMs: GPT-OSS-20B, GPT-OSS-120B, Llama-3.1-8B, Gemma 12b, and Claude Sonnet 4.5 using a rigorous zero-shot evaluation framework across a large dataset of 5,939 patients and seven cancer types. The results of this comparison will clarify where LLMs can reliably replace or augment manual abstraction, where additional safeguards are needed, and which model families appear most promising for future deployment to enhance the efficiency and timeliness of cancer registries.

## Methods

### Data

UNC Health (UNCH) is a large health system with an integrated electronic health record that serves patients across the state of North Carolina (NC). UNCH consists of 16 hospitals and hundreds of clinical practices and has delivered care to millions of patients across NC, the surrounding regions, and from distant referrals. The UNCH hospital-based cancer registry maintains quality standards for Commission on Cancer accreditation as a National Cancer Institute (NCI) designated comprehensive cancer program, thus ensuring high-quality, trustworthy data that can be used as ground-truth data for model evaluation. The UNCH hospital-based cancer registry curates data for individuals that are diagnosed and/or treated for cancer at a subset of UNCH hospitals and practices and reports all cases to the NC Central Cancer Registry (NCCCR), which is a central registry meeting the NAACCR Gold Standard for Registry Certification [23]. For all cases, the UNCH hospital-based cancer registry collects information on patient demographics, diagnosis, first course treatment from all treating facilities, and recurrence. All data is entered in NAACCR format by certified Oncology Data Specialists. Because the UNCH hospital-based cancer registry is directly linked to longitudinal EHR data and contains high-quality, NAACCR-standardized variables curated through manual abstraction, it provides a practical setting to evaluate whether LLMs can support aspects of the cancer registrar workflow.

Figure 1 shows a STROBE diagram of the process to select a cohort with data in the UNCH hospital-based cancer registry. Using a previously established linkage between the UNCH EHR and the NCCCR[24], we selected patients who were diagnosed with cancer between April 1, 2014 and December 31, 2022, where the primary site of the cancer was in the liver, lung, breast, prostate, colon, ovary, or uterus. These cancer sites were chosen based on diversity of common treatment pathways and thus variation in complexity of documentation in the EHR. We then selected only patients who did not have a previous cancer diagnosis in the UNCH hospital-based cancer registry. Finally, we selected only patients who were diagnosed and had their first course of treatment completed at a UNCH hospital-based cancer registry reporting facility as defined in the registry as a Class of Case of 14. Because all patients were diagnosed and had their first course of treatment completed at a single health system, each patient’s reports should include details of all registry variables we want to extract. This allows us to focus on the ability of each LLM to extract values that are in the text, as opposed to inferring based on potentially missing information (i.e. a patient who was diagnosed at UNCH but treated at a different hospital). EHR and clinical report data were extracted from the Carolina Data Warehouse for Health, UNCH’s enterprise clinical data warehouse. The dataset was composed of 5,939 patients with 1,688,432 total clinical reports. Table 1 shows a demographic summary of the cohort. Data were accessed on April 17, 2025. The authors had access to information that identified individual participants due to the free-text nature of clinical reports. Only a subset of authors directly viewed the reports (JTF, MJS, PJL, SN, IWZ).

**Figure 1:**
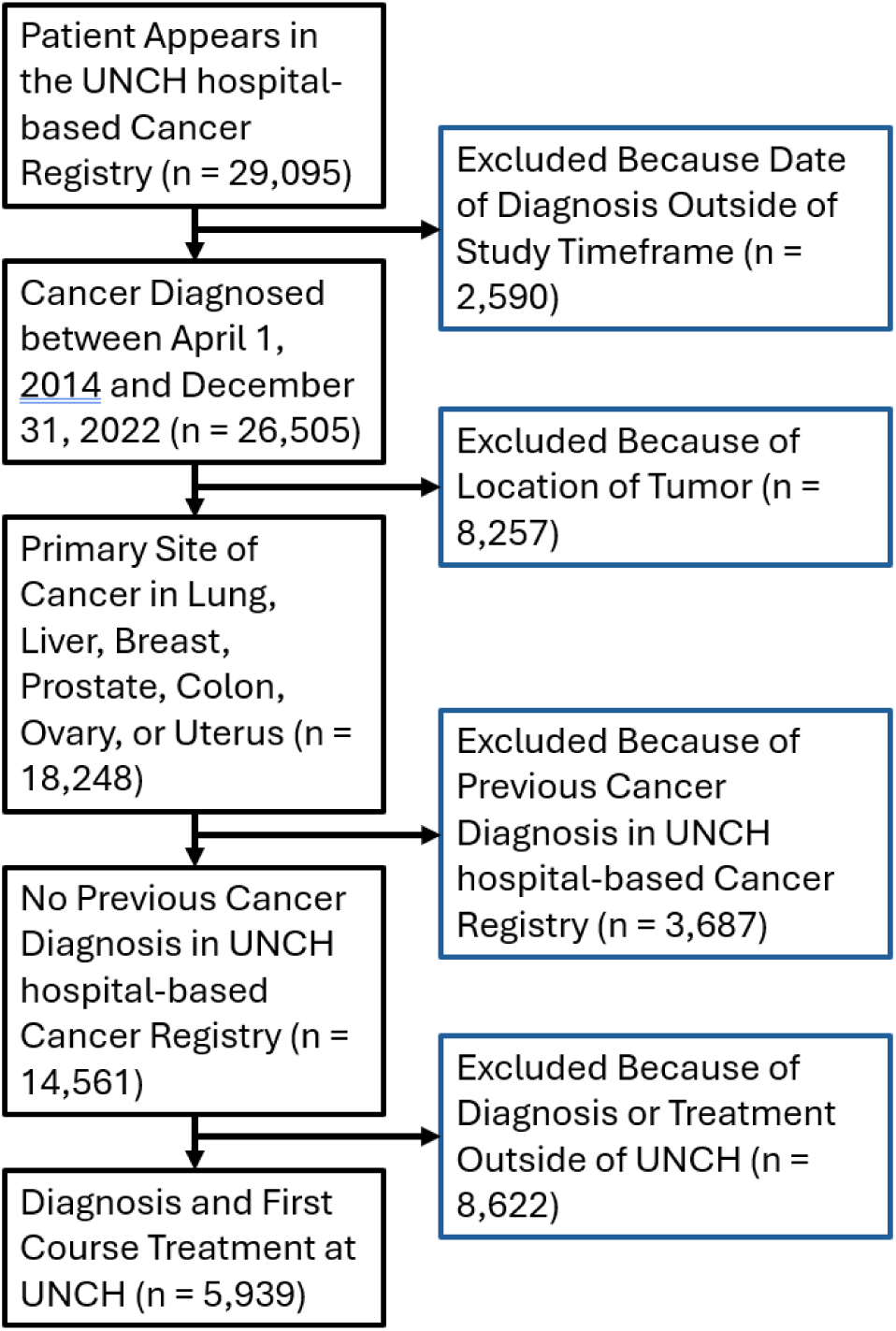
STROBE cohort selection diagram. Selection of patients with inclusion and exclusion criteria.

**Table 1:** Sample Characteristics (N = 5,939)

| Variable Name | Value |
| --- | --- |
| Age (years), mean (SD) | 63.9 (12.3) |
| Age Group, n (%) |  |
| 18-24 | 21 (0.4%) |
| 25-34 | 84 (1.4%) |
| 35-44 | 275 (4.6%) |
| 45-54 | 813 (13.7%) |
| 55-64 | 1740 (29.3%) |
| 65-74 | 1893 (31.9%) |
| 75-84 | 886 (14.9%) |
| 85+ | 227 (3.8%) |
| **Sex, n (%)** |  |
| Female | 3254 (54.8%) |
| Male | 2685 (45.2%) |
| **Race, n (%)** |  |
| White | 4175 (70.3%) |
| Black | 1340 (22.6%) |
| Asian | 91 (1.5%) |
| American Indian or Alaska Native | 50 (0.8%) |
| Other | 283 (4.8%) |
| **Ethnicity, n (%)** |  |
| Hispanic | 219 (3.7%) |
| Non-Hispanic | 5720 (96.3%) |
| **Tumor Primary Site (ICD-O-3 Topographical Code), n (%)** |  |
| Breast (C50) | 1537 (25.9%) |
| Lung (C34) | 1321 (22.2%) |
| Prostate (C61) | 1297 (21.8%) |
| Liver (C22) | 544 (9.2%) |
| Colon (C18) | 538 (9.1%) |
| Uterus (C54 & C55) | 432 (7.3%) |
| Ovary (C56) | 270 (4.6%) |

Cancer patients have many encounters in the health care system, leading to a large number of individual clinical reports. Previous studies have shown a degradation in the ability of LLMs to extract information with very large contexts [25–27]. To partially address this, we limited the set of reports sent to the model by both type and date. We included only the five report types that were determined by two medical oncologists (KRH and WAW) to be most relevant to cancer diagnosis and treatment: Pathology, Radiology, Progress Notes, History and Physical, and Operative Notes. We also limited reports to those that were within 60 days before and 365 days following the date of diagnosis recorded in the UNCH hospital-based cancer registry. These decisions helped the majority of report text fit within the context window for each model while containing the information we aimed to extract. This resulted in 304,199 reports being used for this study.

We selected eight NAACCR variables for extraction and comparison. These cover a range of allowed values (dates, low cardinality, and high cardinality) that allow us to assess model performance across various tasks. These variables are representative of the overall types of NAACCR variables, allowing us to hypothesize about extending this work to a larger set of NAACCR variables. These variables, allowed values, and descriptions are shown in Table 2.

**Table 2:** Set of NAACCR variables extracted for comparison to the UNCH hospital-based registry values. Name, item number, allowed values, and descriptions come directly from the NAACCR Data Dictionary.

| Variable Name | NAACCR Item Number | Number of Allowed Values | Allowed Values | NAACCR Description |
| --- | --- | --- | --- | --- |
| Date of Diagnosis | 390 | Date | Any Date Formatted Like YYYYMMDD | Date of initial diagnosis by a recognized medical practitioner for the tumor being reported whether clinically or microscopically confirmed. |
| Date of 1<sup>st</sup> Course Treatment | 1270 | Date | Any Date Formatted Like YYYYMMDD | Date of initiation of the first therapy for the cancer being reported, using the CoC definition of first course. |
| Laterality | 410 | 7 | 0-5,9 | Code for the side of a paired organ, or the side of the body on which the reportable tumor originated. This applies to the primary site only. |
| Grade | 440 | 9 | 1-9 | Code for the grade or degree of differentiation of the reportable tumor. |
| Summary Stage | 759 | 9 | 0-5, 7-9 | Code for summary stage at the initial diagnosis or treatment of the reportable tumor. |
| Primary Site | 400 | 36 | Four-digit ICD-O-3 Topography Codes (i.e. C34.1) | Code for the primary site of the tumor being reported using ICD-O-3. |
| Regional Nodes Positive | 820 | 95 | 00-90, 95, 97-99 | Records the exact number of regional nodes examined by the pathologist and found to contain metastases. |
| Regional Nodes Examined | 830 | 96 | 00-90, 95-99 | Records the total number of regional lymph nodes that were removed and examined by the pathologist. |

### Modeling

We used five language models: GPT-OSS 20b and 120b [8], LLAMA 3.1 8B Instruct [9], Gemma 12b [28], and Claude Sonnet 4.5. We selected these to cover a range of model sizes and to be able to compare the performance of one commercial model (Claude) to multiple open-source models.

Zero-shot prompting is significant in the context of cancer registry work because many health systems lack the annotation capacity to fine-tune models for every NAACCR variable. To evaluate the zero-shot ability of each model, we engineered a prompt that included the description of each variable and allowed values from the NAACCR Data Dictionary V24 [29], along with coding instructions from the NCI Surveillance, Epidemiology, and End Results (SEER) Program Coding and Staging Manual [30], then asked for that value for each patient. The prompt structure used is provided in Appendix A.

To simplify result parsing and aggregation, models were instructed to call a custom tool to structure the output into a data model with NAACCR variable name, NAACCR variable number, coded value, and an explanation of the decision. Responses from each LLM were cleaned to ensure compliance with accepted values for each variable.

In cases where the model returned invalid responses, the outputs can be highly variable. We applied limited validation checks to correct the most common output errors: use of separators in dates, primary site missing a period, or the model returning the natural language code definition rather than the allowed value itself. These rules were narrowly applied. For example, returning the exact code definition was valid, while returning partial ones was not. Responses that were not allowed values for a given variable were deemed invalid and included as an incorrect response in the performance calculations.

### Analysis

We calculated multiclass classification metrics (accuracy, balanced accuracy, F1, precision, sensitivity, and specificity) for each NAACCR variable with a set number of classes, using the values in the UNCH hospital-based cancer registry for the ground truth. We only analyzed classifications for grade for diagnoses before 2018 because the coding instructions for grade starting in 2018 became site specific, introducing more variability.

For date values, we calculated multiple accuracy scores. The date of diagnosis has been shown to vary based on the data source [31], and the required level of date precision depends on the use case. For example, cancer registrars abstract a single date for both variables investigated here, but clinical trialists care more about the general timeframe of diagnosis and treatment for determining trial eligibility. For that reason, we calculated the exact accuracy, the accuracy within ±7 days, and the accuracy within ±30 days.

Some patients had a total context length that exceeded the allowed amount set by the model. Because the context window for each model is different, this led to a difference in the final number of patients that each model successfully queried. We calculated metrics for each variable only on the subset of patients that had successful queries for all models. This led to a difference in the total number of patients for each NAACCR variable but ensured consistency in comparing results across models.

## Results

### Data Characteristics

Table 1 summarizes the demographics and primary tumor site of the 5,939 patients included in the study. Patients had a mean age of 63.9 with a standard deviation of 12.3 years. Females were slightly more prevalent at 54.8% of the cohort.

The cohort initially had a median clinical report count of 202 with an IQR of 98 – 377. After limiting by type and date, the cohort had a median clinical report count of 39 (IQR 22-70). However, we still dealt with large context windows, with a median token count per patient of 35,925 tokens with an IQR of 19,160 – 61,176.

### Invalid Responses from the LLM

The models occasionally returned values for NAACCR variables that were invalid for three reasons: 1) the returned value was not one of the possible values in the NAACCR data dictionary, 2) the value was improperly formatted and/or unable to be cast to the appropriate type following parsing, or 3) no value was returned.

LLaMA 3.1 8b and Gemma 12b had the most invalid responses, the bulk of which consisted of variations of code or variable definitions. Claude Sonnet 4.5 and the GPT-OSS models were much better at following the allowable code guidelines, typically deviating in small, predictable ways that were caught by the validation checks. The most frequent invalid responses from those models were instances where they returned no value, often with the explanation that there was insufficient information (qualitatively, LLaMA 3.1 8b and Gemma 12b were more likely to guess). For example, in cases where an exact date could not be established, Claude Sonnet 4.5 and the GPT-OSS models typically responded with the most specific timeframe they could deduce (year or month).

The GPT-OSS models also exhibited a tool call failure rate that was not seen among the other models. GPT-OSS-120b failed roughly one out of every four tool calls and GPT-OSS-20b failed approximately one out of every twelve, while the other models only failed a handful of tool calls each. To complete the modelling using GPT-OSS-120B, the prompt was altered such that a JSON object was included at the end of the text response and subsequently parsed using regular expressions. A system prompt leak by GPT-OSS-20B revealed the inclusion of the phrase “you have no access to tools,” which likely accounts for the issue and could have major implications for use of these endpoints in agentic workflows.

### LLM Extraction of Low Cardinality Variables

Three NAACCR variables had a small number of allowed classes (<10): Laterality, Grade, and Summary Stage. Results for each variable by model and metrics are shown in Figure 2. Claude Sonnet 4.5 and GPT-OSS-120b perform very similarly, with F1 scores for each NAACCR variable within 2% of each other. GPT-OSS-20b performed the next best, with F1 scores about 1% and 3% less for Summary Stage and Grade, respectively, and about 20% less for Laterality. Both Gemma 12b and LLaMA 3.1 8b performed significantly worse across all metrics for this set of NAACCR variables. All models show higher specificity values than other metrics. Appendix B shows tables of results for each variable and model.

**Figure 2:**
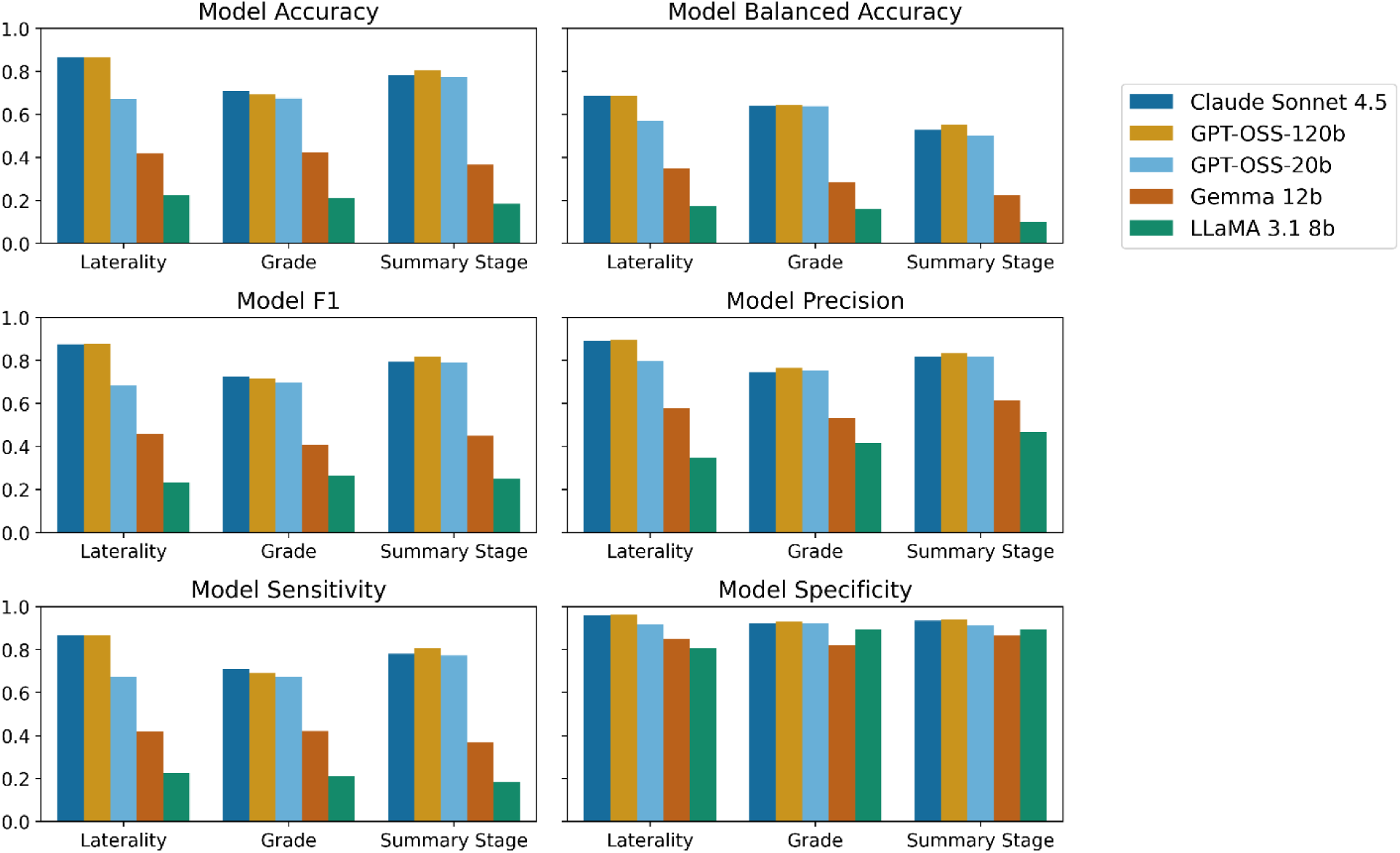
Performance of each model for Laterality, Grade, and Summary Stage. Models are in order by model size. Across all metrics and variables, Claude Sonnet 4.5 and GPT-OSS-120b perform similarly, with GPT-OSS-20b marginally lower. Gemma 12b and LLaMA 3.1 8b perform significantly worse.

As expected for imbalanced datasets, the balanced accuracy scores are consistently lower than the overall accuracy scores. This is because each of these NAACCR variables has 2-3 classes that are minimally represented in the dataset. The high specificity scores indicate that all models are better at indicating what is not in a class.

### LLM Extraction of High Cardinality Variables

Three NAACCR variables had a large number of allowed classes (>30): Primary Site, Regional Nodes Examined, and Regional Nodes Positive. Results for each variable by model and metrics are shown in Figure 3. Appendix B shows tables of results for each variable and model.

**Figure 3:**
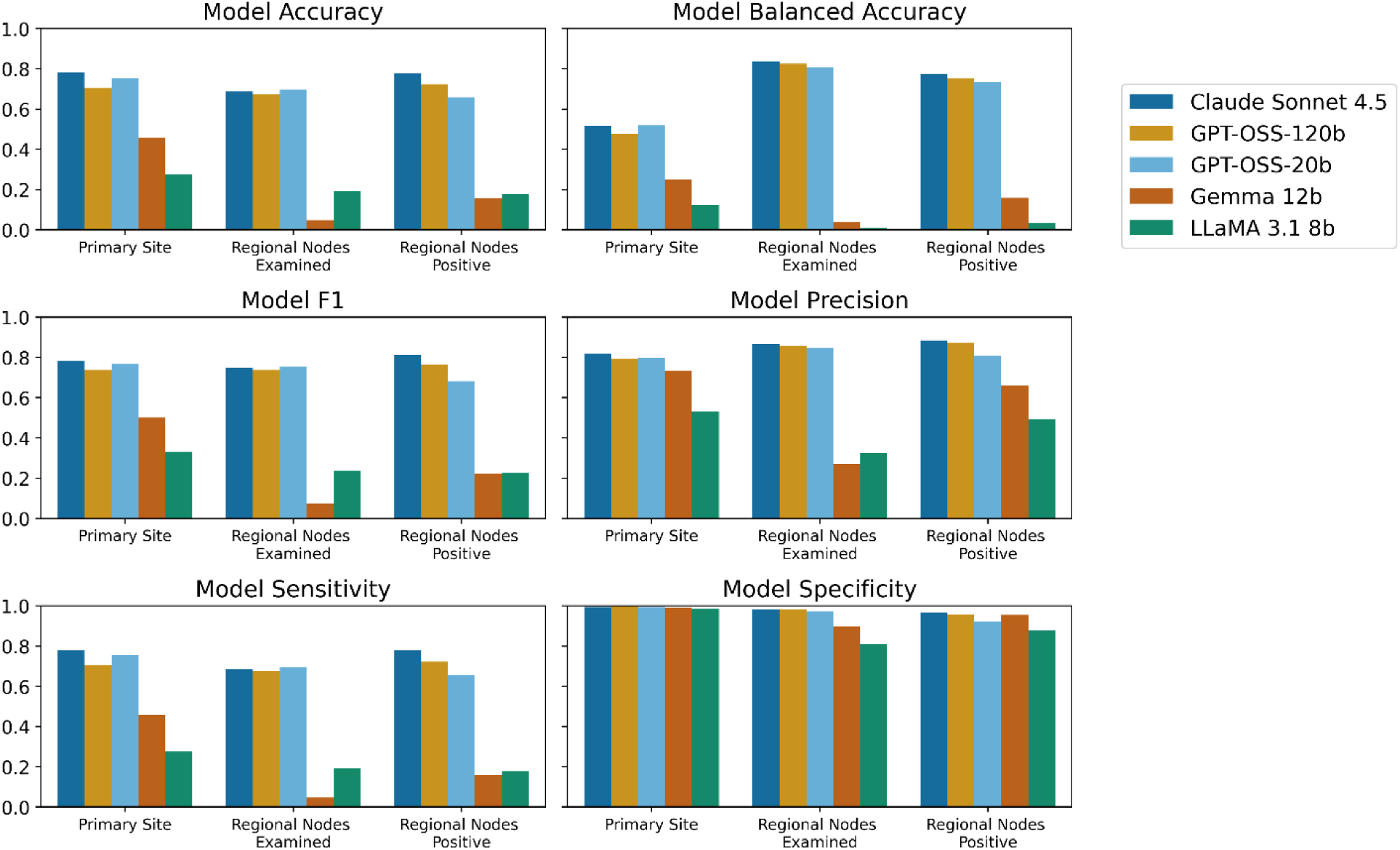
Performance of each model for Primary Site, Regional Nodes Examined, and Regional Nodes Positive. Models are in order by model size. Across all metrics and variables, Claude Sonnet 4.5 and GPT-OSS-120b perform similarly, with GPT-OSS-20b marginally lower. Gemma 12b and LLaMA 3.1 8b perform significantly worse.

Similar to the NAACCR variables with low cardinality, the top performing models were Claude Sonnet 4.5, GPT-OSS-120b, and GPT-OSS-20b. These models had F1 scores within 2% of each other for Regional Nodes Examined. For Primary Site, Claude Sonnet 4.5 and GPT-OSS-20b had F1 scores within 2% of each other with GPT-OSS-120b about 3% lower. For Regional Nodes Positive, Claude Sonnet 4.5 had an F1 score about 5% higher than GPT-OSS-120b, which was about 8% higher than GPT-OSS-20b.

In contrast to the NAACCR variables with low cardinality, GPT-OSS-20b had higher F1 scores than the larger GPT-OSS-120b model on both Primary Site and Regional Nodes Examined.

Gemma 12b performed less well on Primary Site and significantly worse on both Regional Node variables. LLaMA 3.1 8b performed significantly worse on all variables and metrics. All models had higher specificity relative to other metrics.

Extraction of the Primary Site and Regional Nodes Examined have comparable F1 scores despite a very different number of allowed classes (36 vs 96).

### LLM Extraction of Dates

We compared the model performance on the extraction of two dates, the date of diagnosis and the date of first course treatment, and show results in Figure 4. For exact date matches, all models performed poorly, though Claude Sonnet 4.5, GPT-OSS-120b and GPT-OSS-20b still performed significantly better than Gemma 12b and LLaMA 3.1 8b. As expected, the accuracy increased with the larger allowed ranges. Going from requiring an exact match to a match within ±30 days led to a ∼30% improvement for Claude Sonnet and the two GPT-OSS models.

**Figure 4:**
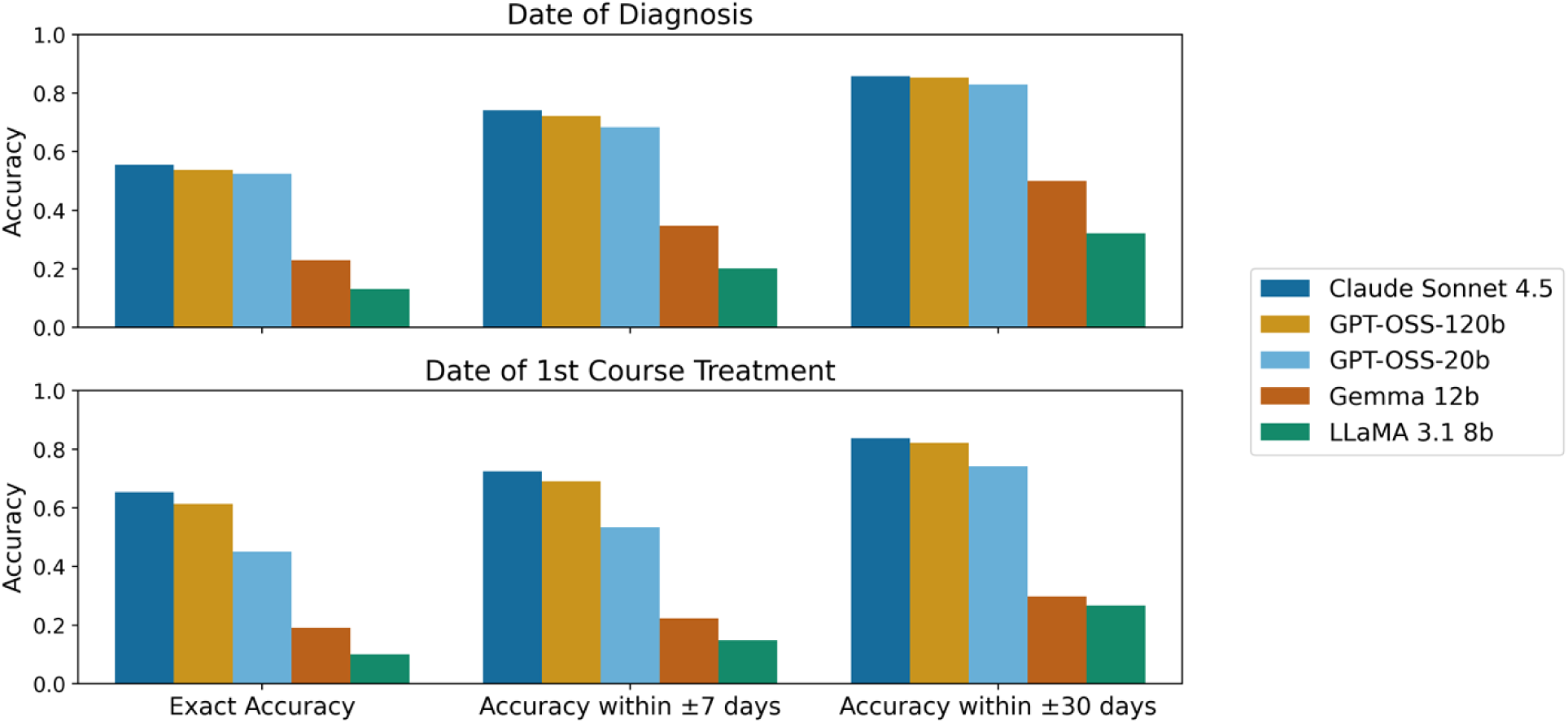
Performance of Date of Diagnosis and Date of 1^st^ Course Treatment extraction by model. Models are in order by model size. We calculate the accuracy of the extraction by exact date, within ±7 days, and within ±30 days.

## Discussion

In this study, we evaluated five state-of-the-art (as of late 2025) LLMs of different sizes on a realistic, high-complexity cancer registry abstracting task involving 5,939 patients, seven tumor types, and eight NAACCR variables. This work demonstrates that LLMs can recover structured cancer registry information from routine clinical documentation with moderate-to-strong performance, but that their utility depends on the specific variable being extracted and the degree of precision required. Ultimately, this study lays the groundwork for building rapid, scalable pipelines for structured cancer information extraction from clinical reports, an important development towards reducing the manual labor required for health care registry data collection, speeding up the availability of these data, and ultimately laying essential work towards natural extensions, such as improving clinical trial recruitment and clinical operations.

We investigated a variety of factors that influence model selection for a real-world clinical registry workflow. One important finding of this work is that model choice matters, but not necessarily in a simple “larger is better” manner. Claude Sonnet 4.5 was the largest and best-performing model overall, but GPT-OSS-120b was often very close in performance and exceeded Claude on Laterality and Summary Stage. This suggests that open-source models may already be viable for some registry workflows, especially in settings where local deployment, data governance, or cost considerations make proprietary models less attractive. At the same time, the consistently weaker performance of Gemma 12b and LLaMA 3.1 8b indicates that not all general-purpose models are equally suited to this task. Generally, our results match recent results showing that open-source LLMs can approach the performance of proprietary models across a range of clinical and biomedical tasks [5,32].

The relatively small performance gap between low-cardinality and high-cardinality variables is also notable. One might expect variables with many possible codes, such as Primary Site or Regional Nodes Examined, to be substantially harder to parse and extract than variables with fewer classes. However, the level of cardinality of the NAACCR variables made only a small difference in the performance of the models. F1 scores for variables with low cardinality were around 0.8 for the best performing models. NAACCR variables with high cardinality had F1 scores averaging around 0.77 for these models. This result is promising when considering real registry workflows with a much larger set of variables of interest that have a large variance in cardinality, indicating the complexity of the mapping from text to coded value does not significantly affect LLM performance.

Date extraction was the most difficult task across all models, though the relative performance of the models was the same. Dates have been previously shown to be challenging to extract from unstructured text [16]. This is likely due to dates commonly being embedded in report metadata rather than in the narrative content itself. In some registry workflows, exact dates are required, but many use cases tolerate some temporal imprecision. The improvement in performance when allowing a broader date window suggests that LLMs may be especially useful for identifying approximate timeframes or candidate dates, even when exact extraction remains unreliable. This has practical implications: systems could be designed to surface likely dates for registrar review rather than attempting fully automated date assignment.

Another important consideration is the interaction between model size, instruction following, valid output generation, and cost. In registry workflows, a model is only useful if it can consistently return a code from a constrained value set and avoid malformed or ambiguous responses. Although the larger models generally produced more reliable outputs, there were still invalid responses and occasional failures to adhere perfectly to the requested format, which matters because even a model with strong nominal classification performance may create substantial downstream burden if its outputs require extensive cleaning or manual correction. At the same time, the cost-performance tradeoff is important: in our computing environment, Claude Sonnet 4.5 was substantially more expensive than the other evaluated models (∼45x), yet its performance gains over GPT-OSS-120b were often modest. Together, these findings suggest that the “best” model for cancer registry abstraction will depend on the deployment context, with some health systems favoring lower-cost open-source models that can be run internally and others prioritizing maximal performance despite higher expense. These tradeoffs are likely to be central to real-world implementation, especially if LLM-based abstraction is scaled across many variables or large case volumes.

These findings indicate that LLMs have significant potential to support aspects of the cancer registry workflow. However, the variability in performance across variable types highlights that fully automated abstraction remains a challenging venture. In practical applications, LLMs may be most effective in a hybrid setting, assisting registrars by pre-populating candidate values or narrowing the search space, rather than fully replacing manual abstraction.

This study has several limitations. Models were evaluated using a zero-shot prompting approach, which may not be as efficient as task-specific optimization approaches such as few-shot prompt engineering, retrieval-augmented generation, or fine-tuning. Additionally, the EHR and registry data were derived from a single health system; as documentation practices and types of cancers seen may differ across institutions, the generalizability of our approach remains untested.

### Future Research

Future research should incorporate other methods to improve extraction accuracy of NAACCR variables, particularly through advanced, targeted prompting strategies, and the integration of retrieval-based methods to better manage long clinical contexts. Additionally, expanding the set of NAACCR variables and investigating results stratified by primary site would provide additional insight into the broader applicability of this work. Researchers could also study whether smaller, clinically focused models might outperform the general-purpose foundation models evaluated in this study while offering lower inference costs. Continued investigation is also required to determine how LLM-based systems can be effectively integrated into clinical data workflows in a manner that supports, rather than replaces, expert human judgment. One such option could be a comparison between LLM-augmented registry abstraction and standard registry abstraction to directly compare time, accuracy, and cost with a goal of enhancing registry efficiency and timeliness while supporting the accuracy appropriate for the clinical application.

This study provides a systematic evaluation of LLM performance on a realistic cancer registry abstraction task and highlights both the potential and current limitations of these models in this domain. Although LLMs are not yet a complete solution for automated registry abstraction, they represent a significant step toward more scalable and efficient clinical data extraction.

## Data Availability

The EHR data used in this study were collected by the Carolina Data Warehouse for Health at the University of North Carolina at Chapel Hill. Free-text clinical records are highly confidential and sensitive and hence cannot be released. Researchers interested in the data used in this study can contact the University of North Carolina at Chapel Hill Office of Human Research Ethics: https://research.unc.edu/human-research-ethics/.

### Appendix A LLM Prompt

Here we show the prompt structure used to extract each variable. The prompt was structured the same for each variable, with {} indicating prompt variables that were updated for each NAACCR variable based on the NAACCR Data Dictionary Version 24. The prompt variables are defined as

- NAACCR_variable_name: NAACCR Variable Name
- NAACCR_ID: NAACCR Data Item Number
- NAACCR_description: Text description of NAACCR variable
- NAACCR_instructions: Any instructions or notes included in the NCI SEER Program Coding and Staging Manual (2024)
- NAACCR_variable_codes: allowed codes with descriptions
- set_of_notes: Concatenated note text for patient

**System Prompt**: You are a helpful assistant to a cancer registrar.

**User Prompt**: Your task is to read a set of clinical notes and extract a NAACCR variable for entry into a cancer registry. Here is the name, ID Number, and a description of the variable:

Variable name: {NAACCR_variable_name} NAACCR ID: {NAACCR_ID}

Description: {NAACCR_description}

The coding instruction for the variable are {NAACCR_instructions}

The possible values for the variable are shown in the following dictionary, where the keys are the output codes and the values are a description of that code:

{NAACCR_variable_codes}

Provide the most appropriate output code for the variable bases on the following notes:

{set_of_notes}

### Appendix B Tables of Results

Here we show numerical results for all NAACCR variables, models, and metrics. We calculated metrics for each variable only on the subset of patients that had successful queries for all models.

#### Date Of Diagnosis (n = 5227)

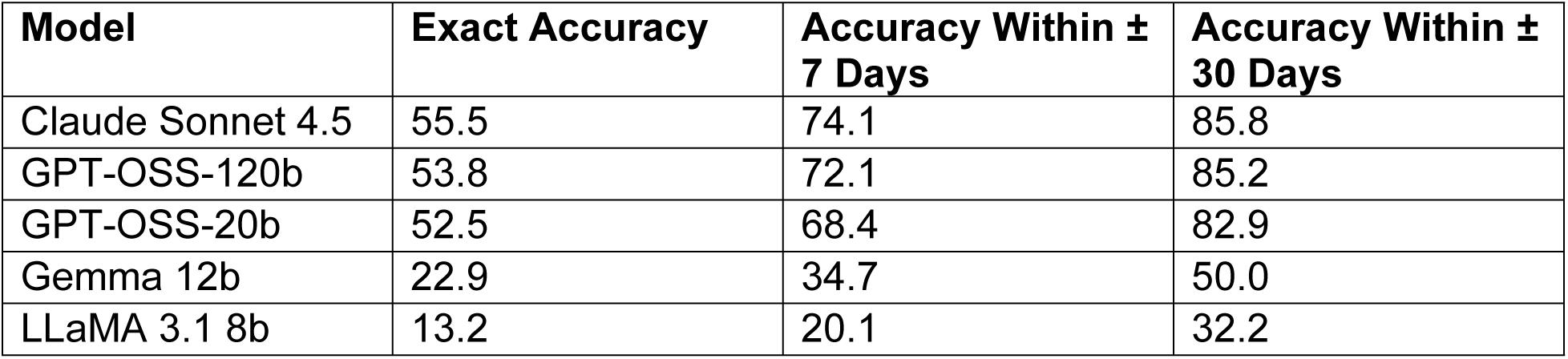

#### Date Of 1^st^ Course Treatment (n = 5200)

| Model | Exact Accuracy | Accuracy Within ± 7 Days | Accuracy Within ± 30 Days |
| --- | --- | --- | --- |
| Claude Sonnet 4.5 | 65.4 | 72.4 | 83.7 |
| GPT-OSS-120b | 61.4 | 69.1 | 82.2 |
| GPT-OSS-20b | 45.0 | 53.4 | 74.2 |
| Gemma 12b | 19.1 | 22.3 | 29.7 |
| LLaMA 3.1 8b | 10.1 | 14.8 | 26.7 |

#### Primary Site (n = 3760)

| Model | Accuracy | Balanced Accuracy | F1 | Precision | Sensitivity | Specificity |
| --- | --- | --- | --- | --- | --- | --- |
| Claude Sonnet 4.5 | 78.1 | 51.7 | 78.4 | 81.9 | 78.1 | 99.3 |
| GPT-OSS-120b | 70.6 | 47.7 | 73.7 | 79.3 | 70.6 | 99.5 |
| GPT-OSS-20b | 75.4 | 51.9 | 76.7 | 79.6 | 75.4 | 99.3 |
| Gemma 12b | 45.8 | 25.0 | 50.0 | 73.2 | 45.8 | 99.1 |
| LLaMA 3.1 8b | 27.5 | 12.3 | 33.2 | 53.1 | 27.5 | 98.4 |

#### Laterality (n = 5227)

| Model | Accuracy | Balanced Accuracy | F1 | Precision | Sensitivity | Specificity |
| --- | --- | --- | --- | --- | --- | --- |
| Claude Sonnet 4.5 | 86.6 | 68.6 | 87.5 | 89.1 | 86.6 | 95.9 |
| GPT-OSS-120b | 86.6 | 68.8 | 87.9 | 89.7 | 86.6 | 96.2 |
| GPT-OSS-20b | 67.3 | 57.2 | 68.3 | 79.8 | 67.3 | 91.8 |
| Gemma 12b | 42.0 | 34.9 | 45.8 | 57.8 | 42.0 | 85.0 |
| LLaMA 3.1 8b | 22.6 | 17.4 | 23.3 | 34.8 | 22.6 | 80.8 |

#### Grade (n = 2837)

| Model | Accuracy | Balanced Accuracy | F1 | Precision | Sensitivity | Specificity |
| --- | --- | --- | --- | --- | --- | --- |
| Claude Sonnet 4.5 | 70.9 | 64.1 | 72.4 | 74.6 | 71.0 | 92.2 |
| GPT-OSS-120b | 69.3 | 64.4 | 71.6 | 76.4 | 69.3 | 92.9 |
| GPT-OSS-20b | 67.4 | 63.8 | 69.8 | 75.2 | 67.4 | 92.4 |
| Gemma 12b | 42.1 | 28.3 | 40.7 | 53.2 | 42.2 | 81.9 |
| LLaMA 3.1 8b | 21.1 | 16.0 | 26.7 | 41.5 | 21.1 | 89.6 |

#### Summary Stage (n = 5202)

| Model | Accuracy | Balanced Accuracy | F1 | Precision | Sensitivity | Specificity |
| --- | --- | --- | --- | --- | --- | --- |
| Claude Sonnet 4.5 | 78.1 | 52.8 | 79.3 | 81.6 | 78.1 | 93.4 |
| GPT-OSS-120b | 80.7 | 55.2 | 81.8 | 83.3 | 80.7 | 93.9 |
| GPT-OSS-20b | 77.2 | 50.3 | 78.8 | 81.9 | 77.2 | 91.2 |
| Gemma 12b | 36.6 | 22.3 | 45.1 | 61.6 | 36.6 | 86.5 |
| LLaMA 3.1 8b | 18.5 | 9.9 | 25.0 | 46.7 | 18.5 | 89.2 |

#### Regional Nodes Positive (n = 5227)

| Model | Accuracy | Balanced Accuracy | F1 | Precision | Sensitivity | Specificity |
| --- | --- | --- | --- | --- | --- | --- |
| Claude Sonnet 4.5 | 77.9 | 77.5 | 81.4 | 88.4 | 77.9 | 96.6 |
| GPT-OSS-120b | 72.2 | 75.1 | 76.2 | 87.4 | 72.2 | 95.5 |
| GPT-OSS-20b | 65.7 | 73.3 | 68.3 | 80.7 | 65.7 | 92.3 |
| Gemma 12b | 15.8 | 16.0 | 22.0 | 65.9 | 15.8 | 95.5 |
| LLaMA 3.1 8b | 17.8 | 3.2 | 22.7 | 49.1 | 17.8 | 87.6 |

#### Regional Nodes Examined (n = 5227)

| Model | Accuracy | Balanced Accuracy | F1 | Precision | Sensitivity | Specificity |
| --- | --- | --- | --- | --- | --- | --- |
| Claude Sonnet 4.5 | 68.7 | 83.8 | 75.0 | 86.9 | 68.7 | 98.2 |
| GPT-OSS-120b | 67.6 | 82.5 | 73.8 | 85.6 | 67.5 | 98.0 |
| GPT-OSS-20b | 69.6 | 80.6 | 75.4 | 84.6 | 69.6 | 97.4 |
| Gemma 12b | 4.7 | 4.0 | 7.3 | 27.3 | 4.7 | 89.8 |
| LLaMA 3.1 8b | 19.2 | 1.1 | 23.8 | 32.6 | 19.2 | 81.1 |

